# Bullying, Psychosis-Like Experiences, and Functional Network Connectivity in Adolescents

**DOI:** 10.64898/2026.02.04.26345538

**Authors:** Pablo Andrés Camazón, Ram Ballem, Jiayu Chen, Zening Fu, Vince Calhoun, Godfrey D. Pearlson, Celso Arango, Covadonga M. Díaz-Caneja, Armin Iraji

## Abstract

Bullying is an adverse childhood experience affecting up to one-third of the global population and linked to psychosis-like experiences (PLEs), which increase the risk of psychotic disorders. This study aimed to investigate the association between the severity and persistence of bullying and PLEs, including non-paranoid, hallucination-like experiences and their distress, and whether multiscale brain functional network connectivity (msFNC) mediates the pathway from bullying to PLEs.

**Methods:** We used data from the ABCD Study, a large, multisite, population-based prospective cohort study following U.S. adolescents. We included adolescents with complete bullying and PLEs assessments at the 2- and 3-year follow-ups (T1: n=10,939, 47% females; T2: n=10,102). We examined the associations between bullying severity and temporal exposure and PLEs using linear mixed-effects models. In a 2-year resting-state fMRI subsample (n=5,280), we used a Neuromark framework to analyze whether msFNC statistically mediated the pathway from bullying to PLEs.

**Results:** Higher PLEs were associated with the presence and severity of bullying (non-bullied vs. mild bullying: d=-0.24, CI:-0.30 to −0.19, p<0.0001; moderate vs. severe bullying: d=-0.62, CI:-0.69 to −0.56, p<0.0001). When bullying ceased, PLEs returned to non-bullied levels, whereas persistence over two years led to greater elevations (d=-0.36, CI:-0.43 to −0.29, p<0.0001), with similar patterns for non-paranoid and hallucination-like experiences and their distress. msFNC in paralimbic, default-mode, central executive, somatomotor, temporoparietal, insulotemporal, and frontal networks statistically mediated the association.

**Conclusions:** Bullying is time- and dose-dependently associated with PLEs. msFNC may provide convergent evidence for distributed neurobiological correlates of the association between bullying exposure and PLEs.

## INTRODUCTION

Bullying, defined as intentional and aggressive behavior perpetrated by peers within a context of power imbalance^1^, is a major public health concern^2–7^. Up to one-third of the global population is exposed to bullying victimization during their lifetime^8,9^, leading to long-term health ^2,3^, economic^7^, and societal^2^ consequences. Evidence suggests that bullying exposure increases the risk of multiple mental health issues, including depression, anxiety, PTSD, and psychosis-like experiences in adolescence^10–13^, the latter of which are linked to a heightened risk of subsequently developing psychotic disorders^14^. However, previous studies have faced methodological limitations related to sample representativeness, study design, and measurement tools^11^. Most have not distinguished between the presence of psychosis-like experiences^10,11^ and their associated distress, despite the latter being more clinically relevant because of its stronger association with later psychotic disorders^15^. To our knowledge, the two largest studies to date found no association between bullying and distressing psychosis-like experiences in a cohort of over 11,000 typically developing adolescents^16^, and there was no association with psychotic disorders in a case-control sample of 11,101 individuals^17^, questioning the potential role of bullying as a risk factor for psychotic disorders^16^. However, previous studies did not differentiate between paranoid and non-paranoid psychosis-like experiences, which may have biased the results, given that questions assessing paranoid ideation and bullying exposure may conceptually overlap in the context of active victimization, and subjects with mainly paranoid experiences may be more likely to label interpersonal experiences as bullying^11^. Additionally, the role of bullying severity and temporal patterns of exposure, both critical for supporting causality, remains unclear. Identifying plausible biological mechanisms is also essential for supporting the causal role of bullying exposure in psychosis-like experiences and informing potential interventions. Bullying exposure has been proposed to negatively affect the developing adolescent brain^18^, which may potentially lead to psychosis-like experiences^14,19,20^. However, this hypothesis has not yet been confirmed. Resting-state fMRI data from large population-based cohorts of healthy adolescents^21^ offer a unique opportunity to investigate how bullying exposure may alter functional connectivity between brain intrinsic networks and examine their potential mediating role in the pathway from bullying exposure to psychosis-like experiences. The default mode network has been shown to partially mediate the association between social victimization (a construct including forms of physical and psychological harm) and PLEs. However, to the best of our knowledge, no studies have analyzed the specific effect of bullying on brain networks and its relationship with psychosis-like experiences using fMRI. We used data from the 2- and 3-year follow-up assessments (T1 and T2, for this study) of the population-based Adolescent Brain Cognitive Development (ABCD) cohort, a longitudinal study of brain development and health in children in the US with more than 11,000 participants^22^. We hypothesized that bullying exposure is associated with psychosis-like experiences in a dose- and time-dependent manner, and that brain functional connectivity mediates the pathway from bullying to psychosis-like experiences. We aimed to (1) analyze the role of bullying exposure severity and temporal patterns in its association with psychosis-like experiences, including non-paranoid and hallucination-like experiences, and distress from these experiences (**Fig.1a-b**); (2) analyze the association between bullying exposure and multiscale functional network connectivity (msFNC) (**Fig.1c**); and (3) conduct an explanatory mediation analysis to estimate the indirect effect of msFNC features on the cross-sectional pathway from bullying exposure to psychosis-like experiences at T1, using the average causal mediation effect (ACME) framework ^23^ (**Fig.1c**).

## METHODS AND MATERIALS

### Participants

This study adhered to STROBE^24^ and AGReMA guidelines^25^. Data were collected between June 2016 and March 2021, from children born between 2005 and 2009 who completed the 2- and 3-year visits (T1 and T2 for this study) of the ABCD Study (release 5.0; https://abcdstudy.org/) (Fig.1). Centralized institutional review board approval was obtained from the University of California, San Diego. Parents or caregivers provided written consent, and the children provided verbal assent to a protocol approved by the institutional review board at each of the 21 data collection sites (https://abcdstudy.org/sites/abcd-sites.html). Sites were geographically distributed across the nation’s four major regions. For each site, the catchment area was defined as all schools within 50 miles of the research institution^26^. Race and ethnicity data were collected via caregiver reports and were included to characterize the representativeness of the sample. Participants were excluded if they met any of the following exclusion criteria: non-English proficiency, general magnetic resonance imaging contraindications, history of a major neurological disorder, traumatic brain injury, extreme premature birth (<28 weeks’ gestational age), diagnosis of schizophrenia, intellectual disability, moderate to severe autism spectrum disorder, or substance abuse disorder^27^.

**Figure 1.**
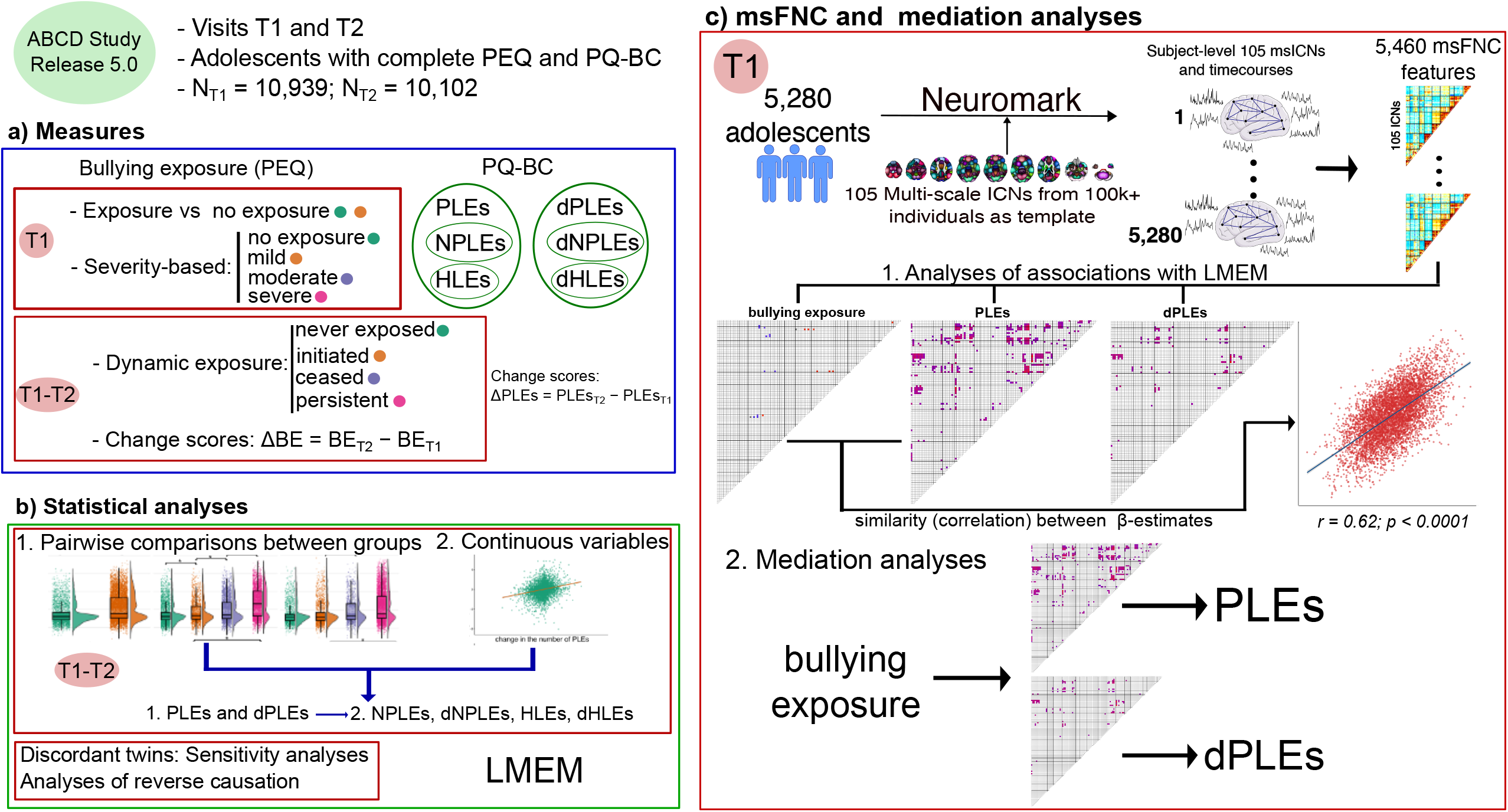
Overview of study design, measures, and analytic workflow. Participants were drawn from ABCD Study (release 5.0), T1, and T2 follow-up visits, including adolescents with complete data on the Peer Experiences Questionnaire (PEQ) and Prodromal Questionnaire–Brief Child Version (PQ-BC). **a)** Measures: bullying exposure (BE) was characterized at T1 by severity-based groups (none, mild, moderate, severe) and longitudinally by dynamic exposure groups (never, ceased, initiated, persistent) between T1 and T2. The number of psychosis-like experiences (PLEs) and associated distress (dPLEs), along with non-paranoid (NPLEs) and hallucination-like experiences (HLEs), were derived from the Prodromal Questionnaire–Brief Child Version (PQ-BC), and change scores were computed. **b)** Statistical analyses: pairwise group comparisons and associations for continuous measures using linear mixed-effects models (LMEMs) adjusted for demographic covariates and family/site clustering. First, we ran the models for PLEs and dPLEs. Second, for significant results, we ran models for NPLEs and HLEs, respectively. Sensitivity analyses were performed in bullying-exposure-discordant twins, and reverse causation was tested by examining whether T1 PLEs predicted subsequent bullying exposure. **c)** Neuroimaging analyses at T1: a Neuromark framework was applied to resting-state fMRI data to identify 105 multiscale intrinsic connectivity networks (msICNs) and derive 5,460 msFNC features for each participant. LMEMs were used to assess the association between msFNC features with bullying exposure and PLEs, and the similarity of brain-wide associations was estimated by correlating β estimates. Mediation analyses tested whether msFNC features statistically mediated the association between bullying exposure and PLEs and dPLEs cross-sectionally at T1.

We used data from the T1 and T2 visits, as these visits had the Peer Experiences Questionnaire (PEQ) available to measure exposure to bullying. We included participants with complete PEQ and Prodromal Questionnaire–Brief Child Version (PQ-BC) data at T1 and T2 visits (n_T1_ = 10,939; n_T2_ = 10,102; **Table 1**, **Fig.1**).

**Table 1.** shows the participants’ demographic characteristics at T1. Continuous variables: mean (standard deviation). Categorical variables: N(%) PLEs: Psychosis-like experiences; NPLEs: Non-paranoid psychosis-like experiences; HLEs: Hallucination-like experiences.

|  | No bullying exposure<br>N = 3,124 | Bullying exposure<br>N = 7,815 |
| --- | --- | --- |
| Demographics |  |  |
| Age (months) | 144 (8) | 144 (8) |
| Sex (female) | 1,462 (47%) | 3,741 (48%) |
| Race |  |  |
| African American | 503 (16%) | 1,132 (15%) |
| Asian | 93 (3%) | 143 (1.8%) |
| Other | 178 (5.7%) | 381 (4.9%) |
| Pacific Islander | 5 (0.2%) | 8 (0.1%) |
| White | 2,345 (75%) | 6,151 (79%) |
| Ethnicity<br>(Hispanic) | 709 (23%) | 1,467 (19%) |
| Annual<br>household<br>income | 8 (3) | 8 (2) |
| Psychosis-like experiences - Prodromal Questionnaire |  |  |
| PLEs | 861 (28%) | 3900 (50%) |
| NPLEs | 830 (27%) | 3,658 (47%) |
| HLEs | 542 (17%) | 2,666 (34%) |
| Bullying exposure - Peer experiences questionnaire |  |  |
| Bullying<br>exposure<br>(continuous<br>variable) | 9 (0) | 14 (4) |

### Measures

We summed the nine peer victimization items of the PEQ as a measure of bullying exposure, each rated on a 1–5 scale (1 = never, 5 = a few times per week) (**supplementary Table 1**). We grouped adolescents according to their exposure to bullying in two different ways: (1) severity-based groups at T1: no exposure (N = 3,124), mild (N = 1,573), moderate (N = 3,316), and severe (N = 2,926) exposure; and (2) dynamic exposure groups based on T1 and T2: never exposed (no bullying exposure at T1 and T2, n = 1,747), ceased exposure (bullying exposure only at T1, N = 1,748), initiated exposure (bullying exposure only at T2, N = 1,109), and persistent exposure (bullying exposure at both time points, N = 5,486) (**Fig.1a**). We used the PQ-BC to assess the number of adolescent-reported psychosis-like experiences and associated distress. Adolescents first answered each question with either yes or no. For each “yes,” adolescents were asked, “Did it bother you?” with numbers ranging from 1–5. Distress scores were calculated as the total number of endorsed questions weighted by the level of distress (i.e., 0=no, 1=yes [but no distress], 2–6=yes [1+score on distress scale]) ^27–29^. Finally, we selected non-paranoid and hallucination-related items to compute non-paranoid and hallucination-like experiences scores, along with their respective distress scores (**supplementary Table 2**).

### Covariates

Covariates included sex, age (in months), race/ethnicity (White, Black, Asian, Native American, Pacific Islander, Hispanic, and other), and annual household income (a 10-level categorical variable).

### Neuroimaging analyses

#### Image acquisition and preprocessing

All subjects were imaged using a 3T scanner (Prisma, Siemens, Munich, Germany); Discovery MR750 (GE Healthcare, Chicago, IL); Achieva dStream or Ingenia CX (Philips Healthcare, Andover, MA) with a 32-channel head coil and completed T1-weighted and T2-weighted structural scans (0.7-mm isotropic)^21^. Participants completed four 5-minute resting-state blood oxygen level–dependent scans with their eyes open and fixated on a crosshair. Resting-state images were acquired in the axial plane using an echo-planar imaging sequence^21^. We downloaded ABCD FastTrack images that passed the consortium’s raw-data quality control (https://nda.nih.gov/). We preprocessed rsfMRI data using a combination of the FMRIB Software Library v6.0 (FSL) toolbox and Statistical Parametric Mapping 12 (SPM) toolbox in the MATLAB R2020b environment. First, we performed rigid body motion correction using the *mcflirt* tool in FSL before distortion correction. We corrected the distortion in the fMRI volume using the *applytopup* tool in FSL 4 based on the field map coefficients obtained with the *topup* tool in FSL^30^. After distortion correction, we discarded 10 initial scans with large signal changes to allow the tissue to reach a steady state of radio-frequency excitation. Next, we warped the fMRI data into the standard Montreal Neurological Institute space based on an echo-planar imaging template and resampled to 3 × 3 × 3 mm^3^ isotropic voxels using the normalization tool in SPM. Finally, we smoothed the resliced fMRI images with a 6 mm full-width at half-maximum Gaussian kernel.

#### Extracting subject-specific multi-scale intrinsic connectivity networks ms(ICNs)

Multivariate-objective optimization independent component analyses with reference (MOO-ICAR) is a spatially constrained independent component analysis approach that estimates intrinsic connectivity networks (ICNs) by optimizing their independence while maintaining their similarity with the reference. We chose MOO-ICAR because it has demonstrated effective performance in capturing subject-specific information while effectively eliminating artifacts^31–33^. We used the GIFT software toolbox (http://trendscenter.org/software/gift) to perform MOO-ICAR, and generate subject-specific ICNs^32,34,35^ with the Neuromark_fMRI_2.2 template (http://trendscenter.org/data) as a reference, which includes highly replicated 105 msICNs across different spatial scales in over 100k individuals^33^.

#### Estimating subject-specific multi-scale functional network connectivity (msFNC)

We ran an additional post hoc cleaning procedure to the msICN time courses to mitigate the effect of the remaining noise, which may not be wholly removed using ICA. The procedure included removing linear, quadratic, and cubic trends, regressing the six motion realignment parameters and their derivatives, replacing outliers with the best estimate using a third-order spline fit to the clean portions of the time courses, and applying bandpass filtering using a fifth-order Butterworth filter with a cutoff frequency of 0.01-0.15 Hz. Subsequently, we computed the subject-specific msFNC by calculating pairwise Pearson correlations between the cleaned msICN time courses. This process resulted in a 105 × 105 symmetric msFNC matrix or a vector of 5,460 FNC features per fMRI scan (19,574 scans), representing the whole-brain functional connectome^36,37^. We further applied Fisher’s Z transformation to improve normality. We computed the mean msFNC of each FNC feature by averaging the scans of the same subject and timepoint. Ultimately, msFNC data were available for 5,280 adolescents.

### Statistical analyses

#### Bullying exposure and psychosis-like experiences associations

We conducted all statistical analyses using R version 4.4.1. We imputed the missing demographic and clinical variables using chained random forests^38,39^. All variables were presented with <2% missing data, except for race (7.74%). Previous analyses of the same sample have shown a small impact of selective attrition on adolescents with and without psychosis-like experiences^40^. We used two analytic approaches to assess the association between bullying exposure and psychosis-like outcomes (**Fig.1a-b**). First, using a categorical approach, we compared psychosis-like outcomes between (1) bullied and non-bullied adolescents cross-sectionally at T1, (2) pair-wise severity-based groups at T1, and (3) pair-wise dynamic exposure groups at T2. Second, we examined the associations between continuous measures of bullying exposure (defined as the summation of all PEQ victimization items) and psychosis-like outcomes, both cross-sectionally at T1 and longitudinally, using change scores (i.e., ΔBE=BE_T2_ − BE_T1_). For both approaches, we ran models for the number of psychosis-like experiences and the associated distress and applied the Bonferroni correction for 38 tests. For significant results after Bonferroni correction, we further examined the associations with non-paranoid and hallucination-like experiences and their respective distress, and applied an additional Bonferroni correction for 68 tests. We applied a log(x+1) transformation to bullying exposure and psychosis outcomes^41^ and z-scored the continuous variables. We used linear mixed-effects models, with family structure nested within sites as a random effect, and the variable of interest and covariates as fixed effects. We computed standardized β*-*estimates with 95% bootstrapped confidence intervals (1,000 iterations) and *R²* for both models, including only the variable of interest and the fully adjusted models. We conducted sensitivity analyses using bullying-discordant twins who typically share a genetic background and environmental risk factors (**Fig.1b**). Finally, to assess reverse causation, we analyzed whether, in non-bullied adolescents in T1, the presence of psychosis-like experiences before T2 (baseline, 1-year, and 2-year visits) increased bullying exposure in T2 compared to adolescents without psychosis-like experiences (**Fig.1b**).

#### Multiscale functional network connectivity and mediation analyses

We used fMRI data from T1, the first time point with available data from the PEQ, PQ-BC, and fMRI (N = 5,280) (**Fig.1c**). First, we used the neuro*combat R package* to adjust for non-biological variance (scanner effects using hashed serial numbers) introduced by multi-scanner MRI and multi-site acquisition protocols in msFNC measurements^42^. We used neuroCombat to adjust msFNC for scanner effects, including head motion, age, sex, race/ethnicity, and household income as covariates to preserve their effects. To analyze associations with msFNC features, we used three sets of linear mixed-effects models with the continuous variable of bullying exposure, the number of psychosis-like experiences, and their associated distress as outcome variables. Each msFNC feature was included as a predictor in one model within each set, using the same fixed and random effects structure of covariates as in previous models, plus head motion (mean frame-wise displacement) and handedness. To assess the similarity between brain-wide associations of psychosis-like outcomes and bullying, we correlated the 5,460 standardized estimates (β-estimates) of bullying exposure with the estimates of psychosis-like outcomes (permutation-based test, N=1000).

Next, we explored whether the msFNC features associated with the number of experiences or related distress statistically mediated the from bullying exposure to these psychosis-like outcomes (path diagram in **Fig.1c**) by estimating their natural indirect effect in the models (average causal mediation effect, ACME). We used standard three-variable path models with bullying exposure as a predictor, the number of psychosis-like experiences and associated distress as outcome variables, and msFNC features as mediators in a counterfactual-based framework^23^. We applied the Benjamini-Hochberg false discovery rate correction (5,460 msFNC).

## RESULTS

### Severity groups

Bullying exposure was associated with a higher number of psychosis-like experiences and distress in a dose-dependent manner (**Fig.2, supplementary Table 3**). At T1, adolescents exposed to bullying had a higher number of experiences (d=-0.60, CI:-0.65 to −0.55, p<0.0001) and more severe distress (d=-0.61, CI:-0.66 to −0.56, p<0.0001) than non-exposed adolescents (**Fig.2a, supplementary Table 3**). A more severe exposure was associated with an incrementally higher number of experiences and distress (no exposure < mild < moderate < severe), with the largest differences between no exposure and severe exposure (number of experiences: d=-1.18, CI:-1.24 to −1.12, p<0.0001; distress: d=-1.21, CI:-1.27 to −1.14, p<0.0001) (**Fig.2a, supplementary Table 4**). The same pattern was observed for non-paranoid and hallucination-like experiences. At T1, bullying exposure (categorical variable) was associated with a higher number and distress from non-paranoid psychosis-like experiences (number, d = −0.55, CI: −0.59 to −0.50; distress: d = −0.55, CI: −0.60 to −0.50) and a higher number and distress from hallucination-like experiences (number, d = −0.44, CI: −0.49 to −0.40; distress: d = −0.43, CI: −0.48 to −0.39). Being in a group with a more severe bullying exposure was also associated with distress from non-paranoid psychosis-like experiences (no exposure vs. severe exposure: d = −1.09, CI: −1.15 to –1.02, p < 0.0001) and hallucination-like experiences (no exposure vs. severe exposure: d = −0.85, CI: −0.91 to –0.80, p < 0.0001) (supplementary **Table 3**). In addition, bullying exposure (continuous variable) was cross-sectionally associated with a higher number of experiences (R²=0.13, CI:0.12 to 0.14, β=0.36, CI:0.34 to 0.38, p<0.0001) and more severe distress (R²=0.14, CI:0.13 to 0.15, β=0.37, CI:0.36 to 0.39, p<0.0001) at T1.

**Figure 2.**
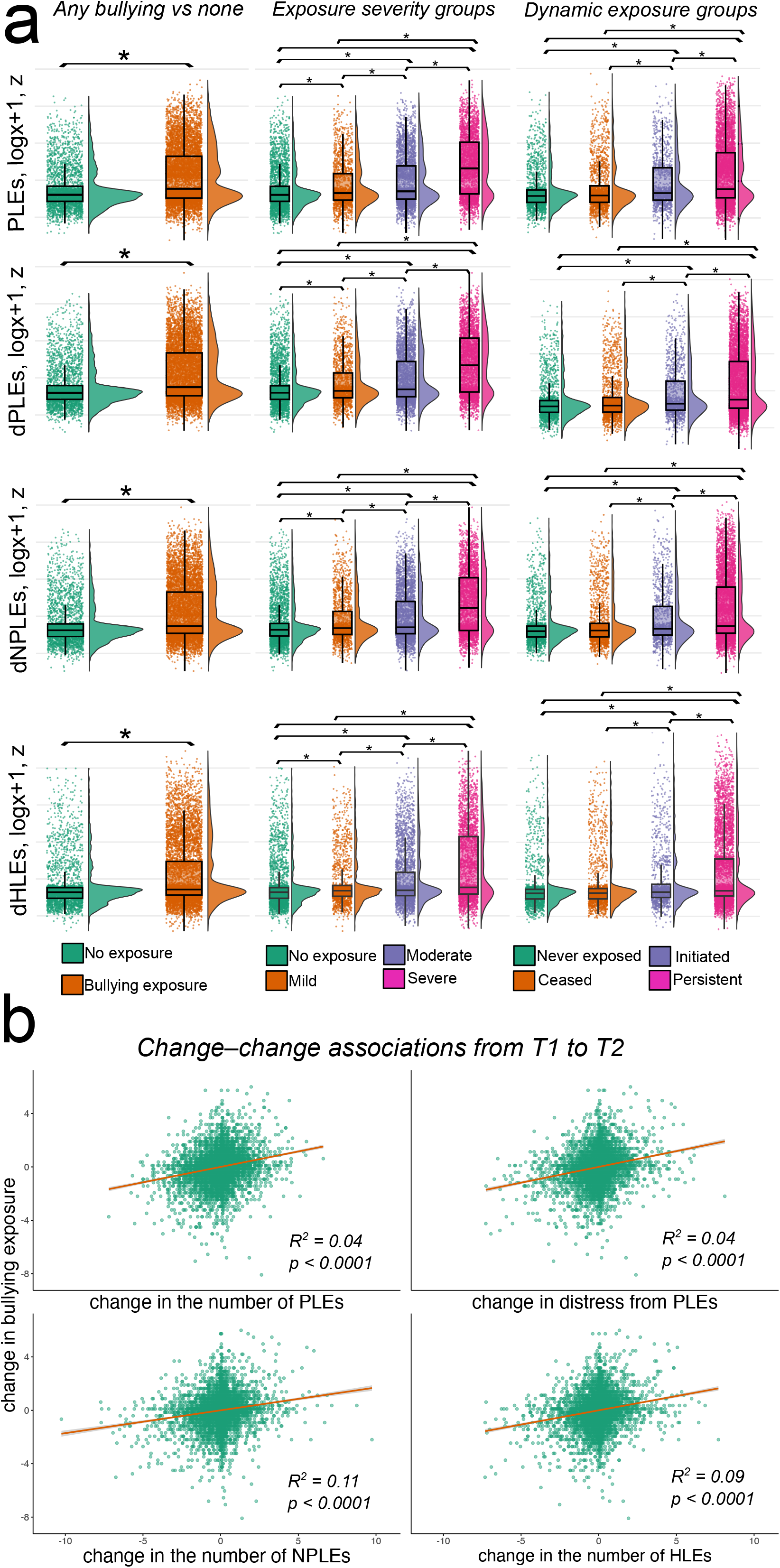
Associations between bullying exposure and psychosis like outcomes. Violin□and□boxplots showing comparisons of psychosis-like outcomes between (i) adolescents with and without bullying exposure cross-sectionally at T1 (first column), (ii) severity-based bullying exposure groups (second column) cross-sectionally at T1, and (iii) psychosis-like outcomes at T2 between dynamic exposure groups (third column). *Statistically significant differences after Bonferroni correction. **b)** Scatterplots showing the association between change in bullying exposure and change in psychosis outcomes from T1 to T2, z-scored. Psychosis-like outcomes were log(x+1)-transformed, z-scored, and residualized using linear mixed-effects models, with family structure nested within the site as a random effect, and sex, age, race/ethnicity, and annual household income as fixed effects. PLEs: number of psychosis-like experiences; NPLEs: number of non-paranoid psychosis-like experiences; HLEs: number of hallucination-like experiences; dPLEs: distress from psychosis-like experiences; dNPLEs: distress from non-paranoid psychosis-like experiences; dHLEs: distress from hallucination-like experiences. T1: first time point, 2-year follow-up; T2: second time point, 3-year follow-up.

### Dynamic groups

Bullying exposure and psychosis-like outcomes were also temporally dependent (**Fig.2, supplementary Table 5**). At T2, the ceased-exposure adolescents showed no significant differences compared with never-exposed adolescents (number: d=-0.13, CI: −0.20 to −0.05, p=0.16; distress: d=-0.12, CI: −0.20 to −0.05, p=0.19). In contrast, if the exposure persisted for two years, the number of experiences and distress were particularly high (initiated exposure vs. persistent exposure, number: d=-0.36, CI: −0.43 to −0.29, p<0.0001; distress: d=-0.38, CI: −0.45 to −0.31, p<0.0001) (**Fig.2a**). The same pattern of dose- and temporal-dependent associations was observed for non-paranoid and hallucination-like experiences (never exposed vs. initiated exposure, number: d = −0.33, CI: −0.41 to –0.24, p < 0.0001; distress: d = −0.31, CI: −0.39 to – 0.22, p < 0.0001; initiated exposure vs. persistently exposed, number: d = −0.32, CI: −0.39 to – 0.25, p < 0.0001; distress: d = −0.34, CI: −0.41 to –0.27, p < 0.0001) and for hallucination-like experiences, though with slightly smaller effect sizes (never exposed vs. initiated exposure, number: d = −0.26, CI: −0.34 to –0.18, p < 0.0001; distress: d = −0.24, CI: −0.32 to –0.16, p < 0.0001; initiated exposure vs. persistently exposed, number: d = −0.24, CI: −0.31 to –0.17, p < 0.0001; distress: d = −0.25, CI: −0.32 to –0.18, p < 0.0001) (**Fig.2a, supplementary Tables 4-5**).

### Longitudinal associations between changes in bullying exposure and changes in psychosis-like outcomes

Additionally, the change in bullying exposure between T1 and T2 was associated with positive changes in the number of experiences (R²=0.04, CI: 0.04 to 0.05, β=0.21, CI: 0.20 to 0.23, p<0.0001) and distress (R²=0.04, CI: 0.04 to 0.05, β=0.21, CI: 0.19 to 0.23, p<0.0001) (**Fig.2b)**. Similar results were observed for non-paranoind and hallucination-like experiences. Changes in bullying exposure were associated with positive changes in the number of non-paranoid psychosis-like experiences (R² = 0.11, CI: 0.10 to 0.12, β = 0.33, CI: 0.32 to 0.35, p < 0.0001) and associated distress (R² = 0.12, CI: 0.10 to 0.13, β = 0.34, CI: 0.32 to 0.35, p < 0.0001) and with the number of hallucination-like experiences (R² = 0.09, CI: 0.07 to 0.10, β = 0.29, CI: 0.27 to 0.31, p < 0.0001) and associated distress (R² = 0.09, CI: 0.07 to 0.10, β = 0.30, CI: 0.28 to 0.31, p < 0.0001).

### Exposure-discordant twins

We found comparable, but weaker, associations in the sensitivity analyses of bullying-discordant twins (no exposure vs. severe exposure, N = 416, number: d = −1.07, CI: −1.31 to –0.84, p < 0.0001; distress: d = −1.07, CI: −1.30 to –0.83, p < 0.0001) (**supplementary Tables S6-S8**). Bullying-discordant twins (never exposed vs. persistently exposed) also showed significant differences, although with smaller effect sizes (number: d = −0.53, CI: −0.71 to –0.35, p < 0.0001; distress: d = −0.53, CI: −0.71 to –0.35, p < 0.0001) (**supplementary Table S7**).

### Psychosis-like experiences as a risk factor for bullying exposure

Among non-bullied adolescents at T1, the presence of psychosis-like experiences at T1 was associated with greater bullying exposure at T2 (N=3,124, d=0.26, CI:0.16 to 0.36, p<0.0001). However, this association was not observed in psychosis-discordant twins (N=148, d=0.10, CI: −0.26 to 0.46, p=0.59).

### Association between multiscale functional network connectivity and bullying exposure

Bullying exposure was associated with hypoconnectivity within the occipitotemporal and paralimbic subdomains, and with hyperconnectivity within the basal ganglia and somatomotor subdomains (**Fig.3**). Effect sizes were small, with the strongest association observed for msFNC between the extrastriate cortex and the left caudate (R²=0.004, CI:0.0001 to 0.015, β=-0.064, CI:-0.092 to −0.038, p=0.016) (**Fig.3d**).

**Figure 3.**
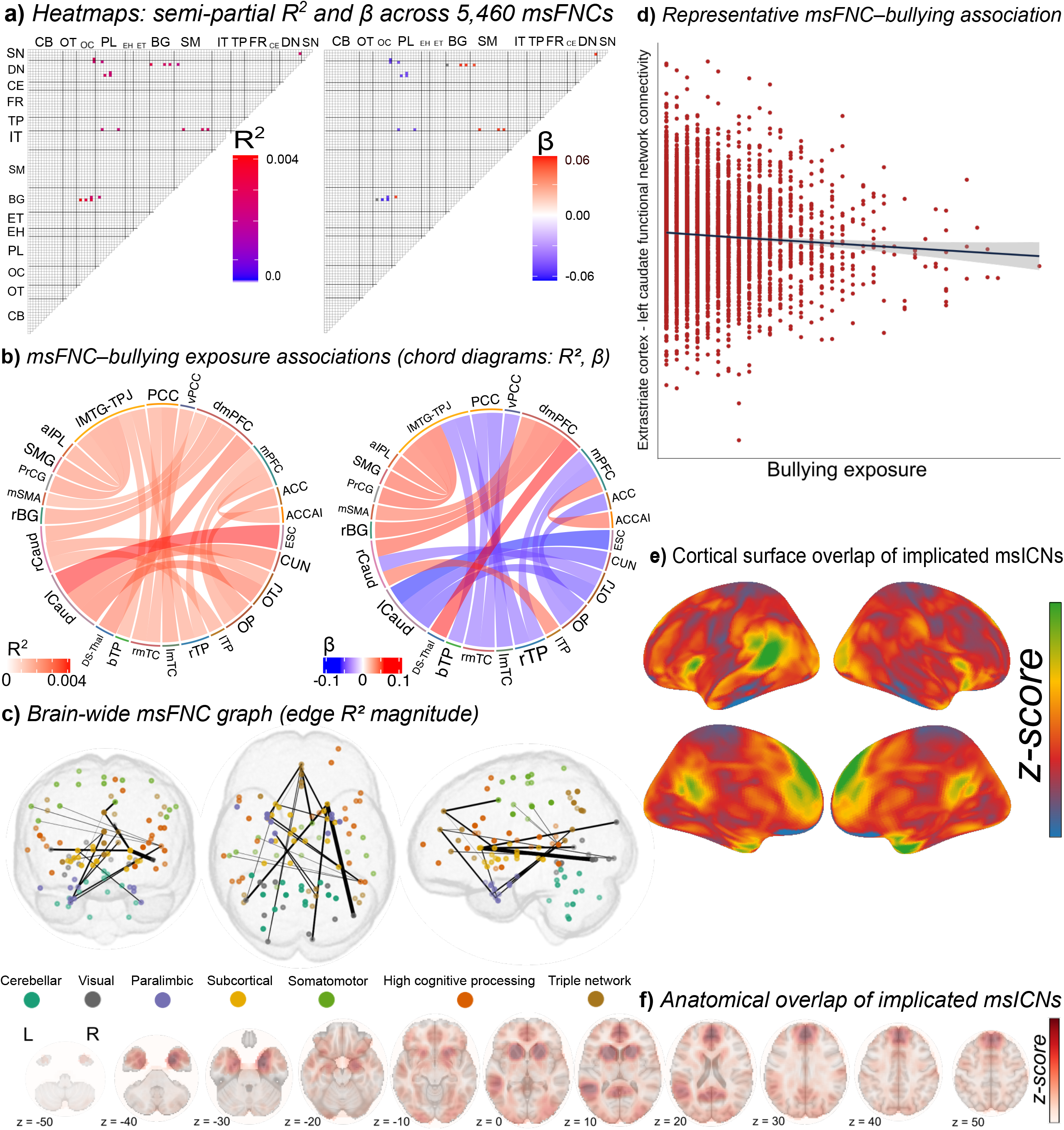
Bullying exposure and multiscale functional network connectivity at T1. **a)** Heatmaps of semi-partial R² (left) and β-estimates (right) for all 5,460 msFNC. Each cell shows one msFNC unique contribution to bullying exposure; colored = significant after false discovery rate correction (p < 0.05), white = nonsignificant. **b)** Chord diagrams of the msFNC–bullying exposure associations (R²; left) and β-estimates (β; right). **c)** 3D MNI152 renders of 105 multiscale intrinsic connectivity networks (msICNs): nodes at peak coordinates; edges weighted by R², line thickness proportional to magnitude; node color denotes functional domain. **d)** Scatter plot showing the association between a representative msFNC feature and bullying exposure. **e)** Cortical surface projections showing anatomical overlap of msICNs with msFNC associated with bullying exposure. **f)** Axial slices (z = −50 to +50 mm) showing anatomical overlap of msICNs with msFNC associated with bullying exposure. R² = semi-partial R-squared; β = standardized regression coefficient; msFNC = multiscale functional network connectivity. CB: Cerebellar; OT: Occipitotemporal; OC: Occipital; PL: Paralimbic; EH: Extended hippocampal; ET: Extended thalamic; BG: Basal ganglia; SM: Somatomotor; IT: Insulotemporal; TP: Temporoparietal; FR: Frontal; CE: Central executive; DN: Default mode network; SN: Salience Network. Full network abbreviations are provided in Supplementary Table 9.

### Association between multiscale functional network connectivity and psychosis-like experiences

The number of psychosis-like experiences was associated with brain-wide msFNC features. Specifically, a higher number of experiences was associated with hypoconnectivity primarily between the cerebellar-frontal, cerebellar-somatomotor, paralimbic-salience, paralimbic-default mode, paralimbic-temporoparietal, insulotemporal-frontal, and insulotemporal-temporoparietal networks, and with increased connectivity primarily observed between the somatomotor-salience, somatomotor-default mode, somatomotor-central executive, somatomotor-frontal, somatomotor-temporoparietal, and frontal-salience networks (**Fig. 4**). Distress from psychosis-like experiences was associated with fewer but similar msFNC features. Distress was associated with hypoconnectivity primarily between the cerebellar-frontal, paralimbic-default-mode, and paralimbic-central executive networks, and with hyperconnectivity primarily between the somatomotor-default mode, somatomotor-central executive, somatomotor-frontal, somatomotor-temporoparietal, insulotemporal-frontal, and insulotemporal-temporoparietal networks (**Fig. 4**). Effect sizes were also small, with the strongest associations involving the left insula and prefrontal cortex for the number of experiences (R² = 0.01, CI: 0.00001 to 0.03, β = 0.08, CI: 0.05 to 0.11, p < 0.001) (**Fig.4**) and with the left inferior frontal gyrus for distress (R² = 0.006, CI: 0.0001 to 0.02, β = 0.08, CI: 0.05 to 0.11, p < 0.001) (**Fig.4**).

**Figure 4.**
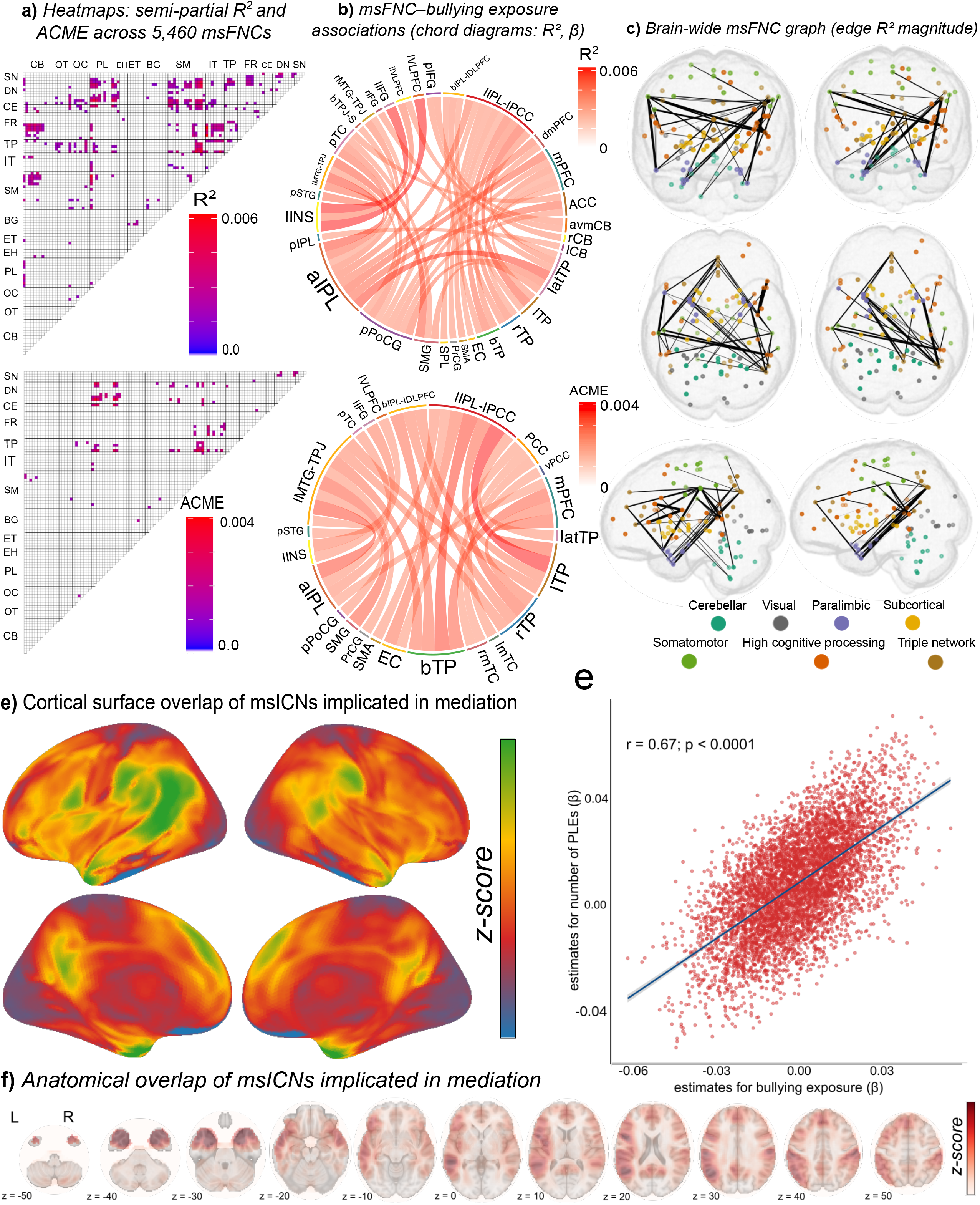
Association between multiscale functional network connectivity and the number and distress of psychosis-like experiences. a, **d)** Heatmaps of β-estimates for all 5,460 msFNC for the number of psychosis-like experiences **(a)** and distress **(d)**. Each cell shows one msFNC unique contribution to the number or distress of psychosis-like experiences; colored = significant after false discovery rate correction (p < 0.05), white = nonsignificant. **b, e)** Chord diagrams of the β-estimates for the number of psychosis-like experiences (b) and distress (e). c) Scatterplot of msFNC between left ventrolateral prefrontal cortex and left insula and the number of psychosis-like experiences; **f)** Scatterplot of msFNC between left inferior frontal gyrus and left insula and distress from psychosis-like experiences. β = standardized regression coefficient; msFNC = multiscale functional network connectivity. CB: Cerebellar; OT: Occipitotemporal; OC: Occipital; PL: Paralimbic; EH: Extended hippocampal; ET: Extended thalamic; BG: Basal ganglia; SM: Somatomotor; IT: Insulotemporal; TP: Temporoparietal; FR: Frontal; CE: Central executive; DN: Default mode network; SN: Salience Network. Full network abbreviations are provided in Supplementary Table 9.

### Mediation analyses

115 msFNC features statistically mediated the pathway from bullying exposure to the number of psychosis-like experiences, primarily between paralimbic-default mode, paralimbic-central executive, paralimbic-temporoparietal, somatomotor-default mode, somatomotor-temporoparietal, and insulotemporal-frontal networks (**Fig.5**).

**Figure 5.**
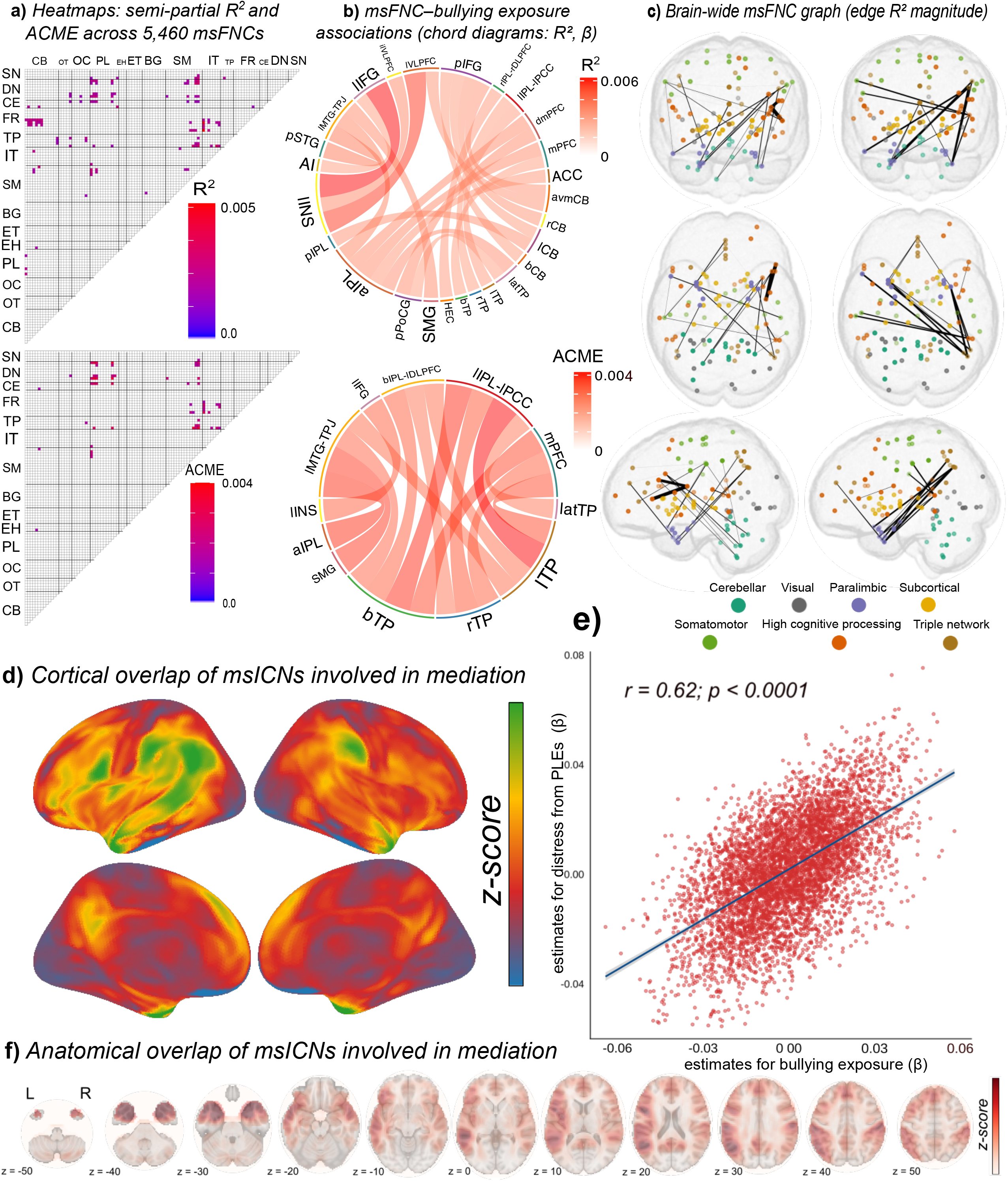
Multiscale functional network connectivity and the association between bullying exposure and the number of psychosis-like experiences. **a)** Heatmaps of semi-partial R² (top) and average causal mediation effect (ACME; bottom) for all 5,460 msFNC. Each cell shows one msFNC unique contribution to the number of psychosis-like experiences (R²) or its mediation of bullying→number of psychosis-like experiences (ACME); colored = significant after false discovery rate correction (p < 0.05), white = nonsignificant. **b)** Chord diagrams of the top 10% msFNC–PLE associations (R²; upper) and top 25% mediation effects (ACME; lower). **c)** 3D MNI152 renders of 105 multiscale intrinsic connectivity networks (msICNs): nodes at peak coordinates; edges weighted by R² (left, top 10%) or ACME (right, top 25%), with line thickness proportional to magnitude; node color denotes functional domain. **d)** Cortical surface projections showing anatomical overlap of mediating msICNs in the bullying→number of psychosis-like experiences mediation. **e)** Scatterplot of msFNC standardized β-estimates for bullying exposure versus standardized β-estimates for the number of psychosis-like experiences; **f)** Axial slices (z = −50 to +50 mm) showing anatomical overlap of mediating msICNs. ACME: average causal mediation effect; PLEs: number of psychosis-like experiences; msFNC: multiscale functional network connectivity. CB: Cerebellar; OT: Occipitotemporal; OC: Occipital; PL: Paralimbic; EH: Extended hippocampal; ET: Extended thalamic; BG: Basal ganglia; SM: Somatomotor; IT: Insulotemporal; TP: Temporoparietal; FR: Frontal; CE: Central executive; DN: Default mode network; SN: Salience Network. Full network abbreviations are provided in Supplementary Table 9.

Similarly, 59 msFNC features statistically mediated the pathway from bullying exposure to distress, primarily between paralimbic-default mode, paralimbic-central executive, somatomotor-default mode, somatomotor-temporoparietal, and insulotemporal-frontal networks (**Fig.6**). ACMEs were small, with msFNC between the left temporal pole and posterior cingulate cortex showing the largest mediation effect for both the number of experiences (ACME=0.004, CI:0.002 to 0.010, p<0.001) (**Fig.5**) and distress (ACME=0.004, CI:0.002 to 0.010, p<0.001) (**Fig.6**). Bullying exposure standardized estimates (β-estimates) of msFNC features were significantly correlated with the estimates from the number of experiences (r=0.67, p<0.0001, **Fig.5**) and distress (r=0.62, p<0.0001, **Fig.6**), suggesting that bullying exposure and psychosis-like outcomes exert comparable effects on whole-brain msFNC patterns.

**Figure 6.** Multiscale functional network connectivity (msFNC) mediates the pathway from bullying exposure to the distress from psychosis-like experiences (dPLE). **a)** Heatmaps of semi-partial R² (top) and average causal mediation effect (ACME; bottom) for all 5,460 msFNC. Each cell shows one msFNC unique contribution to the distress from psychosis-like experiences (R²) or its mediation of bullying→distress from psychosis-like experiences (ACME); colored = significant after false discovery rate correction (p < 0.05), white = nonsignificant. **b)** Chord diagrams of the top 20% msFNC–dPLE associations (R²; upper) and top 25% mediation effects (ACME; lower). **c)** 3D MNI152 renders of 105 multiscale intrinsic connectivity networks (msICNs): nodes at peak coordinates; edges weighted by R² (left, top 20%) or ACME (right, top 25%), with line thickness proportional to magnitude; node color denotes functional domain. **d)** Cortical surface projections showing anatomical overlap of mediating msICNs in the bullying→distress from psychosis-like experiences mediation. **e)** Scatterplot of msFNC β-estimates for bullying exposure versus β-estimates for the distress from psychosis-like experiences; **f)** Axial slices (z = −50 to +50 mm) showing anatomical overlap of mediating msICNs. ACME = average causal mediation effect; dPLEs: distress from psychosis-like experiences; msFNC: multiscale functional network connectivity. CB: Cerebellar; OT: Occipitotemporal; OC: Occipital; PL: Paralimbic; EH: Extended hippocampal; ET: Extended thalamic; BG: Basal ganglia; SM: Somatomotor; IT: Insulotemporal; TP: Temporoparietal; FR: Frontal; CE: Central executive; DN: Default mode network; SN: Salience Network. Full network abbreviations are provided in Supplementary Table 9.

## DISCUSSION

We found that bullying exposure is associated with psychosis-like experiences and their associated distress in a dose- and time-dependent manner in a large sample of typically developing adolescents and in bullying-discordant twins. We identified novel neurobiological correlates linking bullying exposure to the number of psychosis-like experiences and distress through msFNC. Bullying exposure is associated with hypoconnectivity in the occipitotemporal and paralimbic subdomains, and with hyperconnectivity in the basal ganglia and somatomotor subdomains; and functional network connectivity between paralimbic-default mode, paralimbic-central executive, somatomotor-default mode, somatomotor-temporoparietal, and insulotemporal-frontal networks mediate the pathway from bullying to psychosis-like experiences.

The severity and persistence of bullying exposure were linked to a higher number of psychosis-like experiences and distress, whereas cessation of bullying was associated with a return to levels comparable to those of non-bullied adolescents. This reversibility is in line with the observation from Kelleher et al.^11^ using one item on auditory experiences in a smaller sample. Our results confirm and expand it for the associated distress and paranoid and non-paranoid experiences. Given that distressing experiences are associated with a higher risk of psychotic disorders^43^, our findings support the notion that bullying exposure may increase this risk. Contrary to our findings, Martínez et al.^16^ reported no association of peer victimization with distress from psychosis-like experiences, likely due to reliance on a caregiver-reported victimization item, rather than the adolescent-reported PEQ.

The presence of psychosis-like experiences was associated with higher bullying exposure in adolescents not previously victimized, thus suggesting a potential bidirectional relationship. While these adolescents may be more vulnerable to becoming targets, reverse associations likely do not fully account for our findings. As psychosis-like experiences and distress normalize once bullying ceases, it is plausible that anti-bullying interventions drive the reduction in these experiences, rather than spontaneous remission of psychosis-like experiences leading to decreased victimization. Although adolescents with paranoid experiences might be prone to interpret interpersonal interactions as bullying, psychosis-discordant twins without bullying in the previous year did not differ in bullying exposure the following year, suggesting that psychosis-like experiences alone may not be sufficient to report greater bullying exposure. Finally, adolescents with psychosis-like experiences may live in environments with other bullying risk factors. However, we mitigated this possibility by analyzing bullying-discordant twins, who typically share genetic and environmental backgrounds and showed the same associations.

The msFNC between the left temporal pole and the posterior cingulate cortex (PCC) showed the strongest mediation effects for both the number of experiences and distress (**Fig.5** and **6**). These regions are structurally connected, with the PCC acting as a major hub, implicated in internally directed thought and in detecting and responding to salient environmental events that may require behavioral adaptation, partly due to its previous association with schizophrenia. Additionally, a previous study using the Neuromark framework found that anticorrelation between the dorsal PCC and the right postcentral gyrus was associated with early psychosis^46^. Notably, verbal bullying was also linked to increased PCC cortical thickness in patients with an affective first episode of psychosis^47^. The PCC is a key hub of the default mode network (DMN), which has been shown to mediate the association between psychosis-like experiences and social victimization^48^. We showed that not only the DMN but also multiple other networks mediate the pathway from bullying exposure to psychosis-like experiences, including paralimbic, central executive, somatomotor, temporoparietal, insulotemporal, and frontal networks. Prolonged exposure to social defeat—the negative experience of exclusion from the majority group—has been proposed to sensitize the mesolimbic dopamine system, increasing the risk of schizophrenia^49^. Based on this framework, bullying may be more specifically linked to psychosis-like experiences than other forms of trauma unrelated to social exclusion. However, contrary to the original social defeat hypotheses, we did not find that networks within the mesolimbic dopamine system were prominently involved, suggesting that the underlying mechanisms may be more complex and widely distributed^49^. Finally, msFNC estimates from bullying exposure and psychosis-like outcomes were correlated, additionally supporting their link in the developing brain. During adolescence, bullying exposure may alter functional connectivity, contributing to an increase in psychosis-like experiences. Functional alterations may partially normalize when exposure ceases, whereas persistent exposure could exacerbate them, leading to increased psychosis-like experiences; however, this remains speculative given the single time point available for fMRI.

Our study has limitations. First, the PEQ questionnaire was unavailable at visits before the T1, and we could not examine earlier bullying exposure or exclude previously bullied adolescents. Second, mediation analyses were cross-sectional, limiting causal interpretations. Third, the link between psychosis-like experiences and the later development of clinically significant psychotic disorders in the ABCD sample is uncertain. Fourth, we did not adjust for potential confounders such as cannabis use, urbanicity, family environment, other forms of childhood adversity, or other unmeasured confounders in mediation analyses. Fifth, effect sizes were small, likely because of the low signal-to-noise ratio inherent in self-reported measures and fMRI data. Future studies with shorter assessment intervals and more comprehensive data are needed to clarify these associations.

## CONCLUSIONS

Bullying exposure was associated with psychosis-like experiences, including non-paranoid and hallucination-like experiences, and their associated distress in adolescents in a dose- and time-dependent manner, potentially contributing to the development of clinically significant psychotic disorders. Our findings support the implementation of school-based bullying prevention programs^50,51^ and routine screening for bullying in adolescents presenting with psychosis-like experiences and vice versa. Functional network connectivity between paralimbic, central executive, somatomotor, temporoparietal, insulotemporal, and frontal networks may represent distributed neurobiological correlates of this association.

## CODE AVAILABILITY

Preprocessing and data analysis were conducted using MATLAB 9.9.0.1857802 (R2020b) Update 7, alongside the Statistical Parametric Mapping toolbox (SPM 12), the FMRIB Software Library (FSL v6.0), the Group ICA of fMRI Toolbox (GIFT v4.0), RStudio (R version 4.4.1), and Python 3.10.10. MATLAB R2020b can be downloaded from https://www.mathworks.com. The FSL v6.0 toolbox can be downloaded from https://fsl.fmrib.ox.ac.uk/fsl/fslwiki. The SPM 12 toolbox can be downloaded from https://www.fil.ion.ucl.ac.uk/spm/. GIFT v4.0 can be downloaded from https://trendscenter.org/software/gift/. R v4.4.1 can be downloaded from https://cran.r-project.org/. Relevant R packages used included *lme4, partR2, mediation, neuroCombat,* and *emmeans*. These packages can be installed directly from CRAN (https://cran.r-project.org/). Python 3.10.10 can be downloaded from https://www.python.org/downloads/release/python-31010/. The Netplotbrain library used to create brain maps can be downloaded from https://github.com/wiheto/netplotbrain. Additional MATLAB or R scripts used in this study are available from the corresponding authors upon reasonable request.

## DATA AVAILABILITY

The storage and management of the data that support the findings of this study are available from: https://nda.nih.gov/abcd. Access procedures are overseen by the National Institute of Mental Health (NIMH) through the National Data Archive (NDA), visit https://nda.nih.gov/.

## ACKNOWLEDGEMENTS

Pablo Andrés Camazón is supported by the Instituto de Salud Carlos III (ISCIII), Spanish Ministry of Science and Innovation, Río Hortega Program CM24/00125. Covadonga M Díaz-Caneja has received grant support from Instituto de Salud Carlos III, Spanish Ministry of Science and Innovation (PI20/00721, PI23/00625, JR19/00024), and the European Commission (grant 101057182, project Youth-GEMs and grant 101156514, project YOUTHreach). Armin Iraji and Jiayu Chen have received grant support from the National Institutes of Health (R01MH136665). Celso Arango was supported by the Spanish Ministry of Science and Innovation, Instituto de Salud Carlos III (ISCIII) (PI19/01024, PI22/01824), co-financed by the European Regional Development Fund (ERDF), “A way of making Europe”; the European Union – NextGenerationEU (PMP21/00051); CIBERSAM; and the Madrid Regional Government (B2017/BMD-3740, AGES-CM-2), Fundación Familia Alonso, and Fundación Alicia Koplowitz.

## AUTHOR CONTRIBUTION

PAC conceptualized and designed the study, developed the main hypotheses, analytical strategy, and interpreted the results with input from AI, JC and CDC. AI and RB processed the neuroimaging data and implemented and ran the statistical analyses. All authors contributed to the interpretation of the findings. PAC drafted the manuscript and prepared the figures, and all authors critically reviewed and revised the manuscript for important intellectual content. All authors approved the final version of the manuscript.

## REFERENCES

1. Olweus, D. Bullying at School - What We Know and What We Can Do. British Journal of Educational Studies 42, 403–406 (1994).

2. Klomek, A. B., Sourander, A. & Elonheimo, H. Bullying by peers in childhood and effects on psychopathology, suicidality, and criminality in adulthood. The Lancet Psychiatry 2, 930–941 (2015).

3. Arseneault, L. Annual Research Review: The persistent and pervasive impact of being bullied in childhood and adolescence: implications for policy and practice. Journal of Child Psychology and Psychiatry 59, 405–421 (2018).

4. Koyanagi, A. et al. Bullying Victimization and Suicide Attempt Among Adolescents Aged 12–15 Years From 48 Countries. Journal of the American Academy of Child & Adolescent Psychiatry 58, 907–918.e4 (2019).

5. Lereya, S. T., Copeland, W. E., Costello, E. J. & Wolke, D. Adult mental health consequences of peer bullying and maltreatment in childhood: two cohorts in two countries. The Lancet Psychiatry 2, 524–531 (2015).

6. Singham, T. et al. Concurrent and Longitudinal Contribution of Exposure to Bullying in Childhood to Mental Health: The Role of Vulnerability and Resilience. JAMA Psychiatry 74, 1112–1119 (2017).

7. Brimblecombe, N. et al. Long term economic impact associated with childhood bullying victimisation. Social Science & Medicine 208, 134–141 (2018).

8. Modecki, K. L., Minchin, J., Harbaugh, A. G., Guerra, N. G. & Runions, K. C. Bullying Prevalence Across Contexts: A Meta-analysis Measuring Cyber and Traditional Bullying. Journal of Adolescent Health 55, 602–611 (2014).

9. Abregú-Crespo, R. et al. School bullying in children and adolescents with neurodevelopmental and psychiatric conditions: a systematic review and meta-analysis. The Lancet Child & Adolescent Health 8, 122–134 (2024).

10. Catone, G. et al. Bullying victimisation and risk of psychotic phenomena: analyses of British national survey data. The Lancet Psychiatry 2, 618–624 (2015).

11. Kelleher, I. et al. Childhood Trauma and Psychosis in a Prospective Cohort Study: Cause, Effect, and Directionality. AJP 170, 734–741 (2013).

12. Cunningham, T., Hoy,Katrina & and Shannon, C. Does childhood bullying lead to the development of psychotic symptoms? A meta-analysis and review of prospective studies. Psychosis 8, 48–59 (2016).

13. Dam, D. S. van et al. Childhood bullying and the association with psychosis in non-clinical and clinical samples: a review and meta-analysis. Psychological Medicine 42, 2463–2474 (2012).

14. Os, J. van, Linscott, R. J., Myin-Germeys, I., Delespaul, P. & Krabbendam, L. A systematic review and meta-analysis of the psychosis continuum: evidence for a psychosis proneness–persistence–impairment model of psychotic disorder. Psychological Medicine 39, 179–195 (2009).

15. Fusar-Poli, P., Yung, A. R., McGorry, P. & Os, J. van. Lessons learned from the psychosis high-risk state: towards a general staging model of prodromal intervention. Psychological Medicine 44, 17–24 (2014).

16. Martínez, M. et al. Longitudinal study of peer victimization, social support, and mental health during early adolescence. Psychol Med 1–16 (2024) doi:10.1017/S0033291724000035.

17. Bratlien, U. et al. Environmental factors during adolescence associated with later development of psychotic disorders – A nested case-control study. Psychiatry Research 215, 579–585 (2014).

18. Wen, X. et al. Associations of bullying perpetration and peer victimization subtypes with preadolescent’s suicidality, non-suicidal self-injury, neurocognition, and brain development. BMC Med 21, 141 (2023).

19. Arango, C. et al. Preventive strategies for mental health. The Lancet Psychiatry 5, 591–604 (2018).

20. Menken, M. S. et al. Longitudinal alterations in brain morphometry mediated the effects of bullying victimization on cognitive development in preadolescents. Dev Cogn Neurosci 61, 101247 (2023).

21. Casey, B. J. et al. The Adolescent Brain Cognitive Development (ABCD) study: Imaging acquisition across 21 sites. Developmental Cognitive Neuroscience 32, 43– 54 (2018).

22. Volkow, N. D. et al. The conception of the ABCD study: From substance use to a broad NIH collaboration. Developmental Cognitive Neuroscience 32, 4–7 (2018).

23. Tingley, D., Yamamoto, T., Hirose, K., Keele, L. & Imai, K. mediation: R Package for Causal Mediation Analysis. Journal of Statistical Software 59, 1–38 (2014).

24. von Elm, E. et al. The Strengthening the Reporting of Observational Studies in Epidemiology (STROBE) statement: guidelines for reporting observational studies. J Clin Epidemiol 61, 344–349 (2008).

25. Lee, H. et al. A Guideline for Reporting Mediation Analyses of Randomized Trials and Observational Studies: The AGReMA Statement. JAMA 326, 1045–1056 (2021).

26. Barch, D. M. et al. Demographic, physical and mental health assessments in the adolescent brain and cognitive development study: Rationale and description. Developmental Cognitive Neuroscience 32, 55–66 (2018).

27. Karcher, N. R. et al. Assessment of the Prodromal Questionnaire–Brief Child Version for Measurement of Self-reported Psychoticlike Experiences in Childhood. JAMA Psychiatry 75, 853–861 (2018).

28. Cicero, D. C., Krieg, A. & Martin, E. A. Measurement Invariance of the Prodromal Questionnaire–Brief Among White, Asian, Hispanic, and Multiracial Populations. Assessment 26, 294–304 (2019).

29. Loewy, R. L., Pearson, R., Vinogradov, S., Bearden, C. E. & Cannon, T. D. Psychosis risk screening with the Prodromal Questionnaire — Brief Version (PQ-B). Schizophrenia Research 129, 42–46 (2011).

30. Smith, S. M. et al. Advances in functional and structural MR image analysis and implementation as FSL. NeuroImage 23, S208–S219 (2004).

31. Du, Y. et al. Artifact removal in the context of group ICA: A comparison of single-subject and group approaches. Human Brain Mapping 37, 1005–1025 (2016).

32. Du, Y. & Fan, Y. Group information guided ICA for fMRI data analysis. NeuroImage 69, 157–197 (2013).

33. Iraji, A. et al. Identifying canonical and replicable multi-scale intrinsic connectivity networks in 100k+ resting-state fMRI datasets. Hum Brain Mapp 44, 5729–5748 (2023).

34. 34. Du, Y., et al. NeuroMark: An automated and adaptive ICA based pipeline to identify reproducible fMRI markers of brain disorders. NeuroImage: Clinical 28, 102375 (2020).

35. Lin, Q.-H., Liu, J., Zheng, Y.-R., Liang, H. & Calhoun, V. D. Semiblind spatial ICA of fMRI using spatial constraints. Human Brain Mapping 31, 1076–1088 (2010).

36. Allen, E. A. et al. Tracking Whole-Brain Connectivity Dynamics in the Resting State. Cerebral Cortex 24, 663–676 (2014).

37. Jafri, M. J., Pearlson, G. D., Stevens, M. & Calhoun, V. D. A method for functional network connectivity among spatially independent resting-state components in schizophrenia. NeuroImage 39, 1666–1681 (2008).

38. Stekhoven, D. J. & Bühlmann, P. MissForest—non-parametric missing value imputation for mixed-type data. Bioinformatics 28, 112–118 (2012).

39. Buuren, S. van & Groothuis-Oudshoorn, K. mice: Multivariate Imputation by Chained Equations in R. Journal of Statistical Software 45, 1–67 (2011).

40. Osborne, K. J., Barch, D. M., Jackson, J. J. & Karcher, N. R. Psychosis Spectrum Symptoms Before and After Adolescent Cannabis Use Initiation. JAMA Psychiatry 82, 181–190 (2025).

41. Karcher, N. R., O’Brien, K. J., Kandala, S. & Barch, D. M. Resting-State Functional Connectivity and Psychotic-like Experiences in Childhood: Results From the Adolescent Brain Cognitive Development Study. Biological Psychiatry 86, 7–15 (2019).

42. Yu, M. et al. Statistical harmonization corrects site effects in functional connectivity measurements from multi-site fMRI data. Human Brain Mapping 39, 4213–4227 (2018).

43. Wannan, C. M. J. et al. Accelerating Medicines Partnership® Schizophrenia (AMP® SCZ): Rationale and Study Design of the Largest Global Prospective Cohort Study of Clinical High Risk for Psychosis. Schizophr Bull 50, 496–512 (2024).

44. Greicius, M. D., Supekar, K., Menon, V. & Dougherty, R. F. Resting-State Functional Connectivity Reflects Structural Connectivity in the Default Mode Network. Cereb Cortex 19, 72–78 (2009).

45. Leech, R. & Sharp, D. J. The role of the posterior cingulate cortex in cognition and disease. Brain 137, 12–32 (2014).

46. Jensen, K. M. et al. A whole-brain neuromark resting-state fMRI analysis of first-episode and early psychosis: Evidence of aberrant cortical-subcortical-cerebellar functional circuitry. NeuroImage: Clinical 41, 103584 (2024).

47. Fares-Otero, N. E. et al. Triangulating the associations of different types of childhood adversity and first-episode psychosis with cortical thickness across brain regions. Psychol Med 1–14 (2024) doi:10.1017/S0033291724002393.

48. Saxena, A., Liu, S., Handley, E. D. & Dodell-Feder, D. Social victimization, default mode network connectivity, and psychotic-like experiences in adolescents. Schizophrenia Research 264, 462–470 (2024).

49. Selten, J.-P., van der Ven, E., Rutten, B. P. F. & Cantor-Graae, E. The Social Defeat Hypothesis of Schizophrenia: An Update. Schizophr Bull 39, 1180–1186 (2013).

50. Fraguas, D. et al. Assessment of School Anti-Bullying Interventions: A Meta-analysis of Randomized Clinical Trials. JAMA Pediatrics 175, 44–55 (2021).

51. Arango, C., et al. A web-enabled, school-based intervention for bullying prevention (LINKlusive): a cluster randomised trial. eClinicalMedicine 68, (2024).

